# Single-Cell Omics Reveals Dyssynchrony of the Innate and Adaptive Immune System in Progressive COVID-19

**DOI:** 10.1101/2020.07.16.20153437

**Authors:** Avraham Unterman, Tomokazu S. Sumida, Nima Nouri, Xiting Yan, Amy Y. Zhao, Victor Gasque, Jonas C. Schupp, Hiromitsu Asashima, Yunqing Liu, Carlos Cosme, Wenxuan Deng, Ming Chen, Micha Sam Brickman Raredon, Kenneth Hoehn, Guilin Wang, Zuoheng Wang, Giuseppe DeIuliis, Neal G. Ravindra, Ningshan Li, Christopher Castaldi, Patrick Wong, John Fournier, Santos Bermejo, Lokesh Sharma, Arnau Casanovas-Massana, Chantal B.F. Vogels, Anne L. Wyllie, Nathan D. Grubaugh, Anthony Melillo, Hailong Meng, Maksym Minasyan, the Yale IMPACT research team, Laura E. Niklason, Albert I. Ko, Ruth R. Montgomery, Shelli F. Farhadian, Akiko Iwasaki, Albert C. Shaw, David van Dijk, Hongyu Zhao, Steven H. Kleinstein, David A. Hafler, Naftali Kaminski, Charles S. Dela Cruz

**Author notes:** **Corresponding authors** Charles S. Dela Cruz, MD, PhD. Associate Professor, Section of Pulmonary, Critical Care and Sleep Medicine, Department of Internal Medicine & Department of Microbial Pathogenesis, Yale School of Medicine. Director, Center of Pulmonary Infection Research and Treatment (CPIRT). 300 Cedar Street TAC S441-D, New Haven, CT, 06519., Avraham Unterman, MD, MBA. Instructor, Section of Pulmonary, Critical Care and Sleep Medicine, Department of Internal Medicine, Yale School of Medicine, 300 Cedar Street TAC S460, New Haven, CT 06519., Tomokazu S. Sumida, MD, PhD. Assistant Professor, Department of Neurology, Yale School of Medicine, 300 George Street, Room 349, New Haven, CT, 06511. Equal contribution- first authors (Avraham Unterman, Tomokazu S. Sumida). Equal contribution- senior authors (David A. Hafler, Naftali Kaminski, Charles S. Dela Cruz).

## Abstract

A dysregulated immune response against the SARS-CoV-2 virus plays a critical role in severe COVID-19. However, the molecular and cellular mechanisms by which the virus causes lethal immunopathology are poorly understood. Here, we utilize multiomics single-cell analysis to probe dynamic immune responses in patients with stable or progressive manifestations of COVID-19, and assess the effects of tocilizumab, an anti-IL-6 receptor monoclonal antibody. Coordinated profiling of gene expression and cell lineage protein markers reveals a prominent type-1 interferon response across all immune cells, especially in progressive patients. An anti-inflammatory innate immune response and a pre-exhaustion phenotype in activated T cells are hallmarks of progressive disease. Skewed T cell receptor repertoires in CD8+ T cells and uniquely enriched V(D)J sequences are also identified in COVID-19 patients. B cell repertoire and somatic hypermutation analysis are consistent with a primary immune response, with possible contribution from memory B cells. Our in-depth immune profiling reveals dyssynchrony of the innate and adaptive immune interaction in progressive COVID-19, which may contribute to delayed virus clearance and has implications for therapeutic intervention.

## Introduction

SARS-CoV-2, the virus that causes coronavirus disease 2019 (COVID-19), has caused global infection in pandemic proportions already leading to millions of confirmed cases and over half a million deaths worldwide^1^. Infected patients can range from being asymptomatic, to having mild-moderate disease, or more severe disease requiring intensive care unit (ICU)-level care that may include mechanical ventilation and extracorporeal membrane oxygenation (ECMO)^2^. Intensive research efforts are actively ongoing to better understand the pathogenesis and treatment options of this new disease. COVID-19 associated hospitalization data have suggested severe disease disproportionately affects older individuals, those with pre-existing comorbidities, and Black and Hispanic individuals^3^.

There is accumulating evidence to suggest that dysregulated inflammation plays a significant role in the mortality and morbidity of the disease. Patients with severe COVID-19 exhibit substantial immune changes including lymphopenia and increased blood-levels of inflammatory biomarkers such as C-Reactive Protein (CRP), IL-1β, TNF-α, IL-8, and IL-6^4-8^. The magnitude and severity of this inflammatory response have driven significant attention to interventions that modulate immune responses in COVID-19 from corticosteroids to specific cytokine inhibitors^9^. The signaling pathways driven by IL-1β, TNF-α, and IL-6 have been implicated in the pathogenesis of COVID-19^10^ and antibodies against IL-6 receptor have shown early promise^9,11-13^, including our own experience^14^; however, large-scale randomized trials are needed to adequately evaluate their efficacy.

In contrast to early reports emphasizing cytokine storm as a feature of COVID-19, recent studies with deeper profiling of immune cells and with larger cohorts suggest not only a hyper-activated inflammatory response, but also an aberrantly suppressed immune signature^15-17^. These seemingly conflicting results might stem from differences in disease severity and/or from cross-sectional observations at a single time-point that may vary across studies. Given that COVID-19 is an acute viral disease, it is crucial to explore changes in the immune system response across time.

Here, we employed a single-cell multi-omics approach in order to study the dynamics of the innate and adaptive immune system responses in COVID-19, explore the molecular mechanisms that contribute to the progression of the diseases, and assess the effects of tocilizumab, a humanized anti-IL-6 receptor monoclonal antibody. We obtained eighteen paired longitudinal samples of peripheral blood mononuclear cells (PBMCs) from ten COVID-19 patients varying in clinical outcomes and treatment with tocilizumab, as well as 13 samples from age-matched healthy control subjects. We took a comprehensive approach to investigate the immune profiles by using 5’ single-cell RNA sequencing for gene expression (GEX), Cellular Indexing of Transcriptomes and Epitopes by sequencing (CITE-seq)^18^, and B and T cell receptor repertoire analysis. Single-cell results were integrated with clinical and laboratory data, including repeated measurements of SARS-CoV-2 viral load and plasma cytokine levels.

Our results highlight a dynamic type-1 interferon response across all cell types that wanes over time with association to a decrease in viral load and is more prominent in progressive COVID-19 patients. We show that an anti-inflammatory signature in monocytes and a pre-exhaustion phenotype in HLADR^+^CD38^+^ activated T cells are among the hallmarks of progressive disease, with evidence for disrupted connectivity between the innate and adaptive immune system, such as with MHC-class-II to LAG3 signaling. The effects of tocilizumab are different across cell types, possibly related to the wide variability in *IL6R* and *IL6ST* expression. T and B cell receptor repertoire analysis reveal a skewed clonal distribution of CD8 T cells and a primary B cell response with possible contribution from existing memory B cells. Our in-depth immune profiling reveals dyssynchrony of the innate and adaptive immune interaction in progressive COVID-19, which may contribute to delayed virus clearance and has therapeutic implications.

## Results

### Multimodality longitudinal single-cell assessment of PBMCs from COVID-19 patients across severities and time

In order to gain deeper insight into the immune response of COVID-19 patients across disease severities and time course of the disease, we adopted a multimodality single-cell approach to study 18 PBMC samples from 10 patients at various time-points. Age- and sex-matched healthy subjects (n=13), whose samples were collected before the COVID-19 pandemic, were used as controls. Single-cell RNA sequencing (scRNA-seq) was performed using a droplet-based single-cell platform (10x Chromium) ^19^, in order to construct 5’ gene expression libraries, as well as surface protein libraries (CITE-seq^18^), T cell receptor (TCR) libraries and B cell receptor (BCR) libraries (Fig 1C). Following filtration and cleanup (see Methods section, available in our Supplementary Materials), 153,554 cells were included in the scRNA-seq analysis (Fig 1F). In addition, we obtained clinical and laboratory information on all patients, including viral loads and cytokine panels. Our data will be publicly available on GEO and the results can be explored through the COVID-19 Cell Atlas Data Mining Site (www.covidcellatlas.com). This site has a graphical user interface for quick visualization of our scRNA-seq data, which allows users to 1) explore the expression levels of single genes or gene sets of interest across all cell types and 2) conduct comparisons of COVID-19 vs controls, progressive vs stable patients, and early vs late time points across all immune cells in our dataset. We believe that sharing our resources with the broad scientific community with this simple and user-friendly toolkit will facilitate the understanding of COVID-19 immunobiology and ultimately help fight this pandemic.

**Figure 1:**
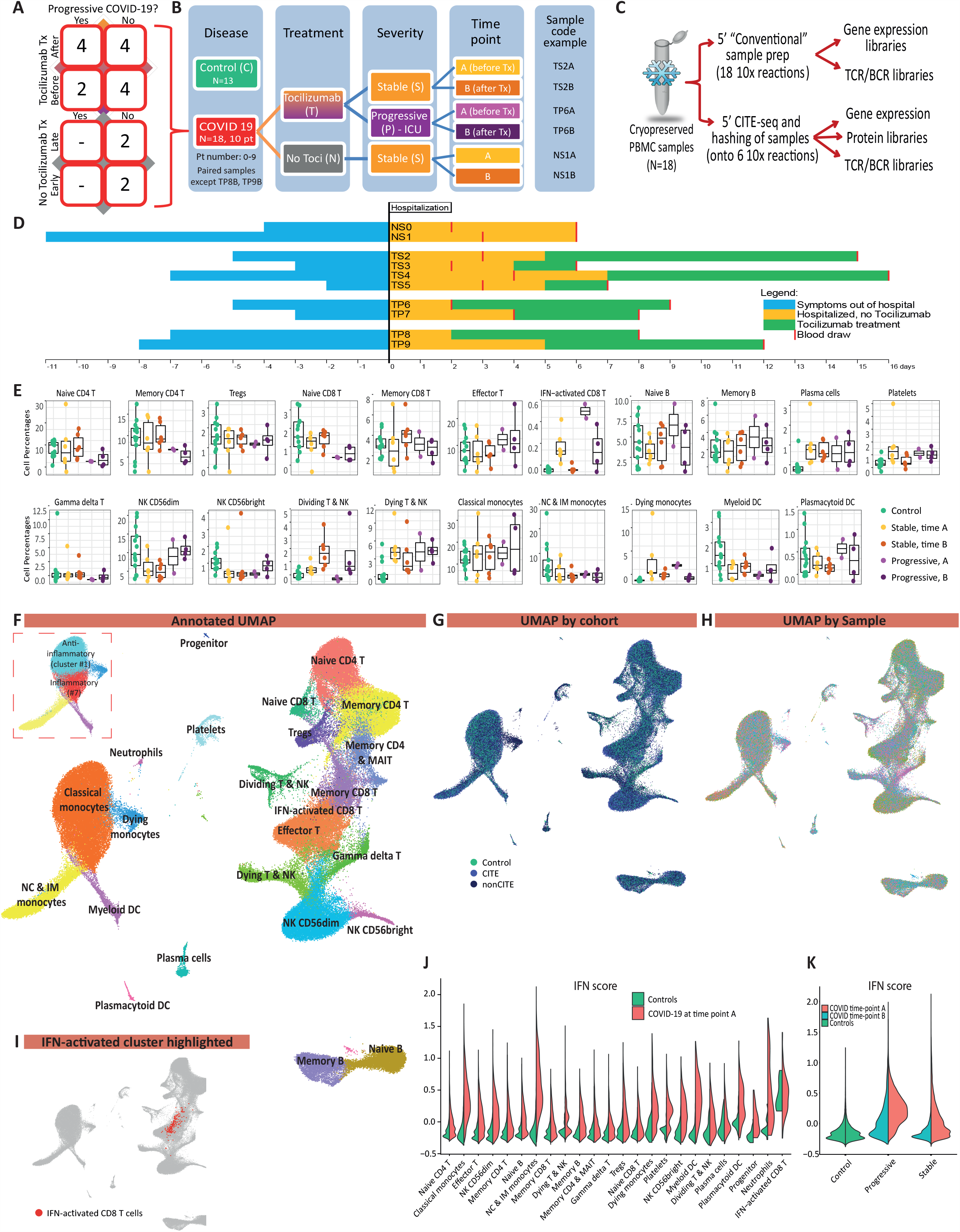
Eighteen PBMC samples from ten COVID-19 patients were included in this study, as well as 13 control samples. All COVID-19 patients had PBMC samples analyzed at two time-points, except for two progressive patients who were only sampled after tocilizumab treatment. **A** Matrix representation of all 18 COVID-19 samples used, according to disease progression, tocilizumab treatment and timing of blood draw. **B** A guide to patient codes and colors used throughout this manuscript. **C** Flowchart of the sample preparation methods and single-cell library types used in this study. Each COVID-19 PBMC sample was split into two after thawing, and processed in parallel by two methods: conventional and CITE-seq. Control PBMC samples were only processed with conventional sample preparation method. **D** Timing of symptoms, hospitalization, blood draws, and tocilizumab treatment for each of the 10 COVID-19 patients. **E** Comparison of differential cell counts (as % of all PBMCs) between patient groups. **F** UMAP embedding of single-cell transcriptomes from 153,554 cells from 18 COVID-19 and 13 control PBMC samples, annotated by cell types. Dashed box shows the two clusters of classical monocytes, an inflammatory cluster (#7) and an anti-inflammatory one (#1). **G** UMAP representation of cells from control samples and COVID-19 samples separated into the two sample preparation methods described in panel C. **H** UMAP by subject, showing relatively little single-subject effects. **I** IFN-activated CD8 T cell cluster highlighted. **J** Interferon score is markedly increased in all cell types in COVID-19 at time point A, relative to controls. **K** Interferon score is higher in progressive vs stable COVID-19 patients, and at time-point A (earlier blood draw) compared to time-point B (later one).

### Patients and samples

Our samples were derived from both stable and progressive COVID-19 patients as part of the Yale COVID-19 IMPACT (Implementing <u>M</u>edical and <u>P</u>ublic Health <u>A</u>ction Against Coronavirus <u>CT</u>) Biorepository. Severe patients (n=4) who required treatment in the ICU and eventually succumbed to the disease were defined as having “progressive” disease, while “stable” disease defined patients (n=6) hospitalized in internal medicine wards and eventually discharged. As part of the COVID-19 treatment algorithm used in the Yale New Haven Health System, patients who required ≥3L/min O_2_ or ≥2L/min but with CRP >70 were treated with tocilizumab. We analyzed PBMCs from two separate blood samples for each patient, an early (A) and a late (B) time-point, except for two progressive patients (TP8, TP9) for whom only a single sample was available (Fig 1A, 1B, 1D). Eighty percent of subjects (8/10) were treated with tocilizumab, with the time-point A sample obtained before and the time-point B sample after the initiation of the treatment. Baseline characteristics (Supplementary Table ST1) including age and sex were similar for both control and COVID-19 patients, while individuals of European ancestry were more prevalent in the controls. Progressive patients did not differ from the stable group with regard to baseline characteristics and timelines (Fig 1D, Supp Table ST1), but all progressive patients expired while all stable patients recovered and were discharged from the hospital.

### Single-cell clustering, cell type annotation, and relative cell counts

Applying Louvain clustering to the filtered and integrated Seurat object^20^, and plotting in uniform manifold approximation and projection (UMAP)^21^ space, 22 cell types were identified and manually annotated (Fig 1F, Supp Fig S1) across 30 cell clusters (Supp Fig S2), with good overlap between different samples (Fig 1I). Automated annotation using SingleR package^22^ (Supp Fig S3) supported the results of the manual annotation. Good overlap was also noted between cells processed with and without CITE-seq (i.e. non-CITE, Fig 1G), except for the dying monocytes cluster which was reduced in CITE samples, possibly due to the exclusion of these dying cells during the lengthier handling times to process CITE-seq samples. Importantly, viability was similar (around 85-90%) for CITE and non-CITE samples before loading the cells to the 10x Chromium Chip. A detailed comparison between the CITE and non-CITE samples (Supp Fig S4) showed high similarity of gene expression, and data sets were therefore combined together for subsequent analysis.

Relative cell counts based on the manual annotation revealed marked differences between control, stable, and progressive samples at the two time-points (Fig 1E). Some notable differences are a relative increase in effector T cells and a decrease in naïve T cells in progressive patients compared to stable ones, as well as an increase in plasmablasts and dividing T & NK cells in COVID-19 patients vs controls, expected during an adaptive immune response to the virus. Cells belonging to the interferon (IFN)-activated CD8 T cell cluster (Fig 1F & 1H), a small cluster of 191 cells characterized by very high expression of IFN stimulated genes (ISGs), are found almost exclusively in COVID-19 patients (p=0.006), especially at time point A.

SARS-CoV-2 RNA was not detected in any of our PBMC samples, concordant with previous reports about the low prevalence of virus in blood samples^7^. In addition, we did not detect expression of *ACE2*, the functional host receptor for SARS-CoV-2^23^, which may diminish the likelihood of PBMC infection.

### Dominant type-1 IFN signature characterizes differentially expressed genes in COVID-19

Type-1 IFN response is elevated in COVID-19 across all cell types (Fig 1J), especially at time-point A, and more so in progressive subjects (Fig 1K, Fig 3A-3C), with the IFN-activated CD8 T cell cluster representing the extreme state. Conventional ISGs, such as *IFI6, IFI44L, LY6E*, and *ISG15*, are markedly increased in COVID-19 patients compared to healthy controls across all major cell types in PBMCs (Fig 3E, Supp Fig S5). Of note, this strong type-1 IFN response is attenuated at time point B in all stable patients but prolonged in some progressive patients (Figure 3A).

Amphiregulin (AREG), a ligand for epidermal growth factor receptor (EGFR) not known as a major IFN stimulated gene, is barely detectable in healthy control PBMCs but is significantly increased in COVID-19 patients’ monocytes, CD4 T cells, NK cells, neutrophils, and DCs (Supp Fig S6). Although AREG is known to play important roles in wound repair and resolution of inflammation^24^, its expression has also been reported to be increased in viral infections of the lung^25^ and induces severe lung pathology in a mouse model of SARS-CoV infection^26^. Type-1 IFN signaling plays an important role in AREG induction within myeloid cells in mice^27^. A recent report using bulk RNA-sequencing showed an increase of AREG in the PBMCs of COVID-19 patients^28^, supporting a potential role of AREG in SARS-CoV-2 induced lung pathology.

Major histocompatibility complex (MHC) class II molecules are decreased in antigen-presenting cells (APCs) of COVID-19 patients compared to controls, especially in progressive patients (Fig 2K, Supp Fig S7). The classical monocytes population sub-cluster into two distinct populations (dashed box in Fig 1F) with different MHC class II (MHC-II) expression (Supp Fig S8): one with low MHC-II expression (cluster #1), subsequently referred to as the anti-inflammatory monocyte, and another with high MHC-II expression representing the inflammatory monocyte (cluster #7). The relative counts for this anti-inflammatory monocyte cluster are higher for COVID-19 patients compared to the controls (Supp Fig S8). *FCGR3A* is significantly downregulated in monocytes in COVID-19 patients compared to healthy controls, in both stable and progressive patients (Fig 3E). This is in line with the decrease in non-classical monocytes in COVID-19, observed in our dataset (Fig 1E, Supp Fig S8) as well as by others^16,29^.

**Figure 2:**
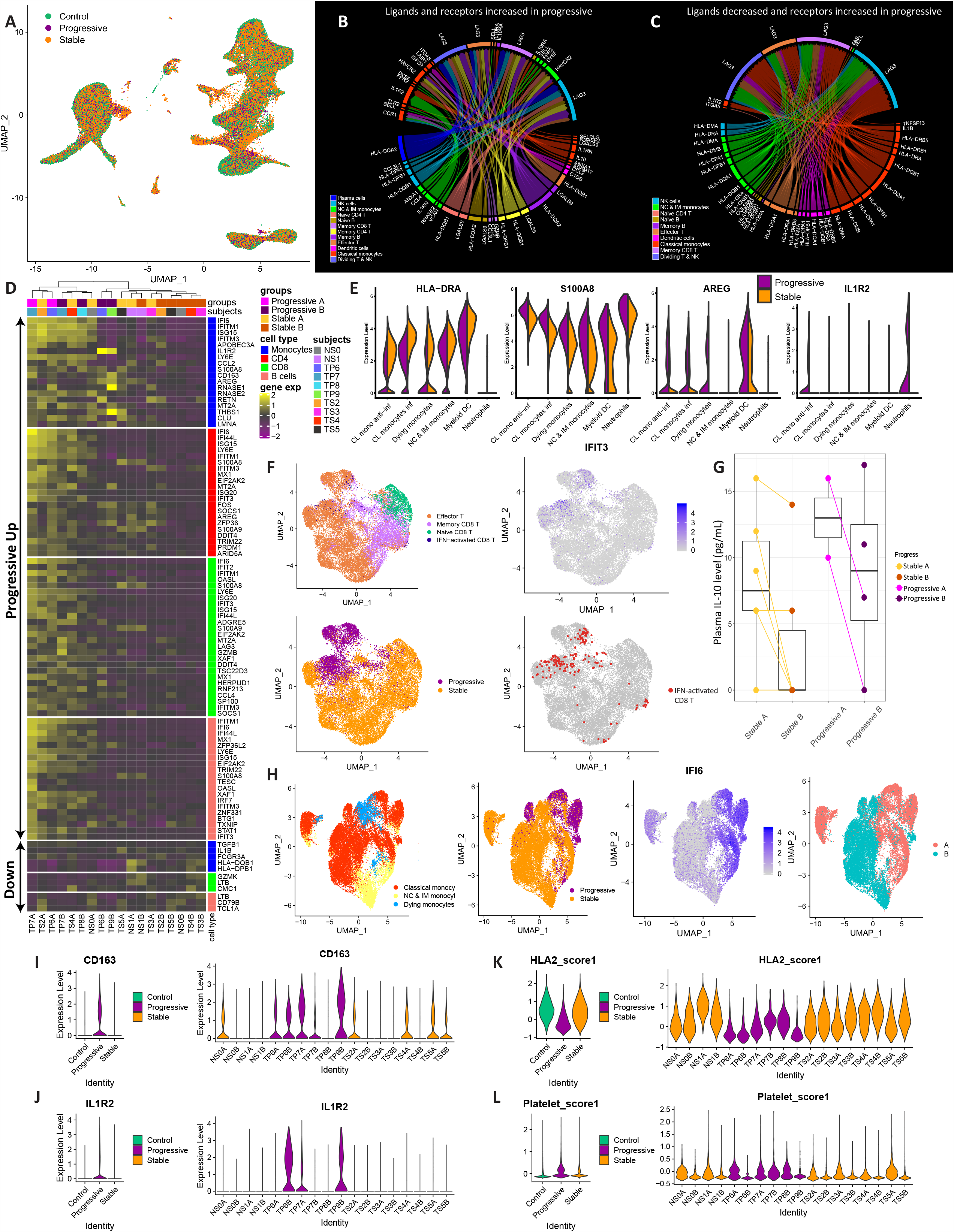
Differences between progressive COVID-19 patients (i.e. severe patients who required admission to the ICU and eventually succumbed to the disease) and stable patients (milder patients hospitalized in internal medicine wards and eventually discharged). **A** UMAP representation of cells from control, stable and progressive subjects. **B** Differential connectivity map (“connectome”) of ligands (bottom half) and receptors (upper half) that are both increased in progressive vs stable patients. **C** Differential connectivity map of ligands that are decreased in progressive patients and receptors that are increased in progressive patients. **D** Heatmap showing the top differentially expressed genes (logFC > 0.5, significant adjusted P values) for each major cell type, comparing stable and progressive patients. Hierarchical clustering separates most of the progressive samples from the stable ones (except for some stable samples at time point A, which cluster with the progressive ones). **E** Violin plots of the expression of *HLA-DRA, S100A8, AREG* and *IL1R2* in myeloid cell clusters, comparing stable to progressive patients. **F** Sub-clustering of CD8 T cells, showing clear separation of cells from progressive individuals, marked by a higher ISG signature (IFIT3 is given as a representative example for ISGs). Most of the cells belonging to the IFN-activated CD8 T cluster are located in the high IFN/progressive pole. **G** Boxplot showing plasma levels of IL-10 (pg/mL) at time-points A and B, corresponding to those of the 18 PBMC samples. Paired samples are connected with a line. IL-10 level is higher in progressive subjects in each time-point, and it decreases from time-point A to B in most subjects. **H** Sub-clustering of all monocytes, showing the pooling of cells from progressive patients to the high ISG area. Note also the marked difference between time point A (earlier blood draw) to time point B (later one), as portrayed further in Figure 3. **I-J** Anti-inflammatory monocyte markers (CD163, IL1R2) are increased in progressive COVID-19. **K** A composite score of all HLA type 2 genes is decreased in progressive relative to stable patients and controls. **L** A composite score of 6 specific platelet genes is increased in progressive vs stable monocytes.

**Figure 3:**
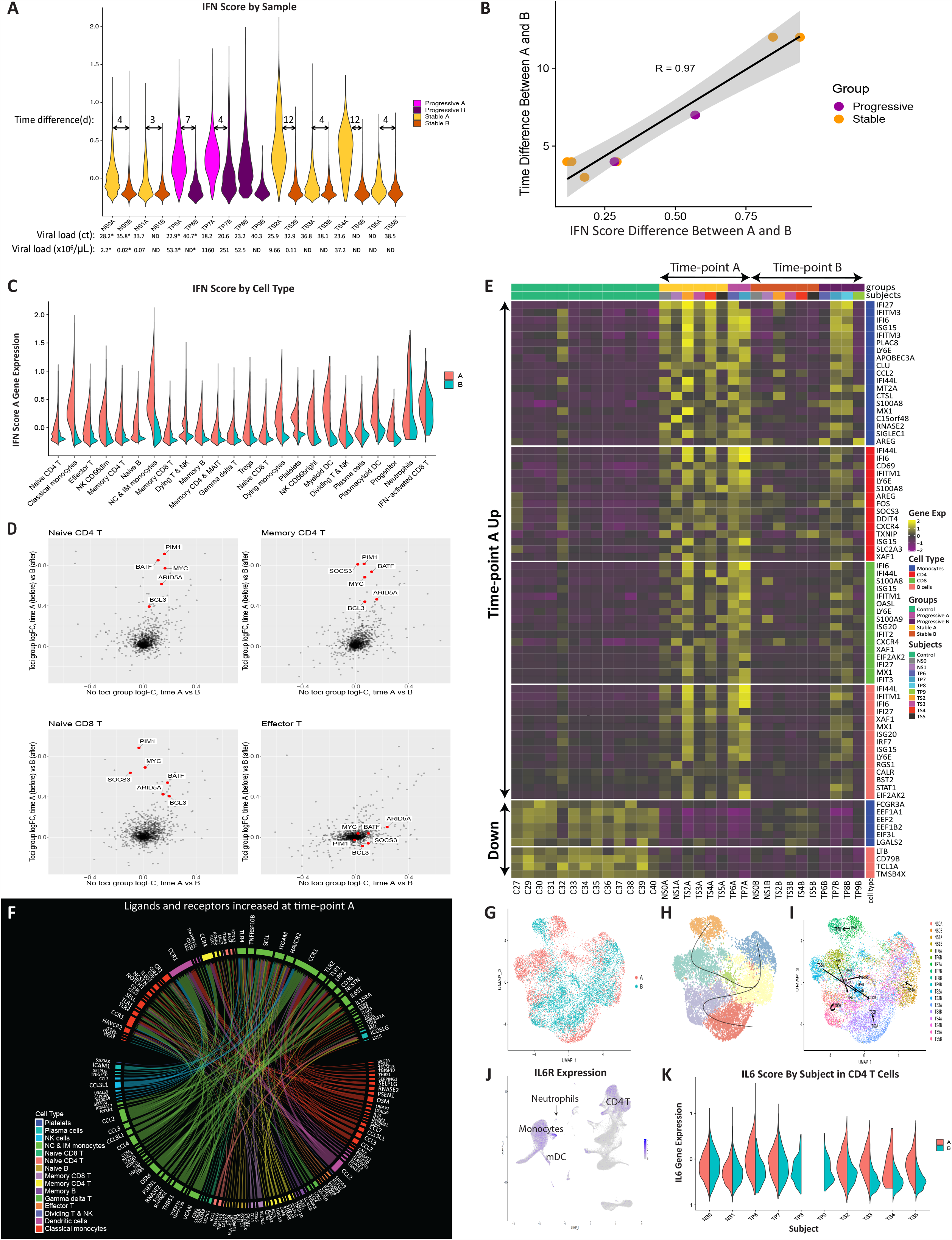
Analysis of changes in gene expression over time in COVID-19 patients, and the effect of anti-IL-6 treatment with tocilizumab. **A** Interferon (IFN) score for each sample in association with the viral load (obtained by nasopharyngeal swabs, except for those marked with * which were obtained by saliva samples). Arrows mark the time difference (in days) between paired samples (i.e. from the same patient) at two time-points: A (early/before tocilizumab treatment) and B (late/after tocilizumab). **B** Scatter plot for the 8 paired samples, showing a very high correlation between the time difference from sample A to B and the respective change in IFN score (scaled) during that time. **C** Interferon score decreases from time-point A to B in nearly all cell types. **D** A scatter plot of the logFC from time-point A to B in patients treated compared to those not treated with tocilizumab. This comparative model demonstrates a marked effect of tocilizumab on IL-6 pathway genes (shown in red) in CD4 and naive CD8 T cells but not in effector CD8 T cells, in which *IL6R* expression is low (see Supp Fig S13 for the full panel with all cell types). **E** Heatmap showing the top differentially expressed genes (logFC > 0.5, significant adjusted P values) for each major cell type, comparing time-point A and B. The level of expression of these genes in control samples is shown as well. **F** Differential connectivity map of ligands (bottom half) and receptors (upper half) that are both increased in time point A compared to time-point B. **G, H, I** Sub-clustering of CD8 T Cells (also refer to figure 2F), with emphasis on time: **G**-UMAP showing differences between time-point A and B on the UMAP space, **H**-pseudotime trajectories diverge towards the two poles of effector T cells at time-point A: one enriched with progressive samples and the other with stable ones, **I**-arrows represent the shift on a UMAP space from time point A to B for each subject, with a general trend of moving from the periphery to the center, except for subject TP7 which maintains a high IFN score (and high viral loads) at time-point B. **J** *IL6R* expression in PBMCs is highest for monocytes, dendritic cells, CD4 T cells (including T regs) and naïve CD8 T cells. **K** IL-6 score in CD4 T cells is decreased in time-point B in all the patients that were treated with tocilizumab (except for TP7), but not in the non-treated patients (NS0, NS1).

In the following sections, we focus on gene expression changes that characterize disease severity and dynamics over the time course of the disease.

### Marked gene expression changes, both stimulatory and suppressive, differentiate progressive from stable patients

We observed marked gene expression differences between stable and progressive patients that span across all cell lineages (Fig 2A-D, Supp Table ST2-ST5). The expression of ISGs is increased in all cell types in progressive subjects, which strongly correlates with their higher viral loads (Fig 3A; correlation coefficient between log10 of viral load and IFN score is 0.8). Interestingly, there is an increased expression of the suppressive cytokine *IL10* in myeloid cells and several additional cell types in the progressive patients (Supp Fig S9). Levels of IL-10 in plasma are known to be increased in severe COVID-19, as reported in our recent study^30^ as well as by others^7,31^. Plasma IL-10 levels for our ten patients show a similar trend (Fig 2G) although not statistically significant as may be expected in this small sample size. Type-1 IFN has been reported to induce IL-10 expression, thus limiting immune-related tissue damage in certain conditions^32-36^. Similar to ISGs, the level of plasma IL-10 decreases from time-point A to B (Fig 2G), and there is a positive correlation (R=0.50, Supp Fig S9) between the IFN score in PBMCs and plasma IL-10, supporting an association between the strength of the IFN response and the suppressive IL-10 response observed in COVID-19 patients.

In addition, we observed a decrease in MHC-II transcripts in antigen-presenting cells (APCs) of progressive subjects (Fig 2C, 2K). Of note, MHC-II expression in APCs of stable patients was more similar to that of control subjects (Fig 2K). Increased IL-10 is known to downregulate the expression of MHC-II^37,38^, possibly explaining this observed decrease in progressive subjects. Of note, a signature of platelets is increased in progressive vs stable monocytes (Fig 2L), suggesting a higher frequency of platelet-monocyte complex formation in progressive patients. Of interest, activated platelets have previously been shown to enhance IL-10 secretion by monocytes^39^.

Together, this suppressive signature of increased IL-10 and decreased MHC-II in progressive patients might serve as a double-edged sword: on the one hand decreasing inflammation and protecting tissues from immune-related damage and on the other hand hampering the ability to mount an effective anti-viral response, as will be further discussed in the next section.

### Anti-inflammatory monocytes characterize progressive patients, despite having higher viral loads

In order to better understand gene expression changes in specific cell types of interest, we sub-clustered monocytes (Fig 2H) and CD8 T cells (Fig 2F). We excluded control subjects before re-clustering to provide better contrast of the transcriptional differences between stable and progressive COVID-19.

Sub-clustering of the monocyte population yielded 23,701 monocytes in seven clusters (Fig 2H, Supp Fig S10). We identified a clear separation between cells of stable and progressive patients, which is driven in part by increased expression of ISGs (e.g. *IFI6, IFITM1, ISG15, IFITM3, LY6E, ISG20, IFI44L*) in progressive patients (Fig 2D, Supp Table ST2). Anti-inflammatory and tissue repair genes are increased in progressive vs stable monocytes, including *CD163, IL1R2, AREG, MRC1*, the co-inhibitory receptor *HAVCR2* (encoding TIM-3) and its ligand *LGALS9* (encoding Galectin-9), and the suppressor cytokine *IL10* (Fig 2B, 2D, 2E, 2I, 2J). The expression of *RNASE2*, encoding a protein with anti-viral activity (mainly against single-stranded RNA viruses)^40,41^, is also increased in progressive patients.

MHC-II molecules were decreased in progressive monocytes as detailed above (Fig 2C, 2D, 2E, 2K). The alarmins S100A8/S100A9 are ranked among the top DEGs increased in progressive vs stable monocytes, which is also observed in SARS-CoV^42^ infection. Given that S100A9 is a marker of myeloid-derived suppressive cells (MDSCs)^43,44^ and can promote IL-10 production and suppressive capacity of MDSCs^45,46^, the anti-inflammatory phenotype of monocytes in progressive patients somewhat resembles that of MDSCs^47^. Indeed, one of the two classical monocyte clusters (cluster #1, Fig 1F-dashed box) is enriched with MDSCs associated genes (*S100A8, S100A9, IL1R2, IL10*) with low expression of MHC-II. Furthermore, this anti-inflammatory monocyte cluster exhibits highly overlapped transcriptional features with a recently identified monocyte population in severe sepsis^48^. Unexpectedly, pro-inflammatory monocyte markers such as *IL1B* and *TNF* are downregulated in COVID-19 monocytes relative to controls (Supp Fig S11), both in progressive and in stable patients, although *IL1B* was slightly less downregulated in stable patients. This observation is consistent with recent reports highlighting an immunosuppressive phenotype in severe respiratory failure in COVID-19 patients^49^.

Taken together, these findings revealed a skewed anti-inflammatory signature of monocytes in progressive patients, which resembles immunoparalysis ^50^. Given that many of these genes associated with an immunosuppressive phenotype are regulated downstream of type-1 IFN signaling (*AREG, IL1R2, S100A8, S100A9, IL10*), this shift of classical monocytes toward MDSC-like suppressive cells might stem from the strong type-1 IFN response. In the context of the higher viral load and a stronger type-1 IFN response observed in progressive patients (Fig 1L, 3A), this shift to a resolution phase --potentially prematurely-- might interrupt appropriate anti-viral immune responses, contributing to deleterious clinical manifestations in severe COVID-19.

### CD8 T cells exhibit an enhanced effector phenotype with higher ISG expression in progressive patients

A detailed analysis of sub-clustered 19,458 CD8 T cells (Figure 2F, Supp Fig S12) shows a clear separation between cells of stable and progressive patients visualized in UMAP space, driven mainly by a higher expression of ISGs in the progressive patients (Fig 2D, 2F), but also by higher markers of effector T cell activation such as GZMB. Effector T cells, a cluster comprised mainly of CD8 T cells (ratio of CD8 to CD4 cells in this cluster is about 6:1), is relatively increased while naïve CD8 T cells cluster is relatively decreased in progressive subjects, also indicative of effector T cell activation and expansion in the CD8 compartment (Fig 1E). Most of the cells from the IFN-activated CD8 T cell cluster are located in the progressive pole and overlap with the effector T cell cluster (Fig 2F). There are clear shifts in the gene expression profile from the early time-point A to the late time-point B (Fig 3G, 3H, 3J) that are mainly driven by the decrease in ISG signature (Fig 3E). The differential connectivity map analysis (Fig 2B, 3C) demonstrates an increased expression of the co-inhibitory receptor *LAG3* in T lymphocytes of progressive patients, while their ligands, which are MHC-II molecules, are decreased in antigen presenting cells (APCs).

### Gene expression changes over time suggest slower virus clearance in some progressive patients

The time course of COVID-19 disease is characterized by significant shifts in many genes (Fig 3E) and ligand-receptor interactions (Fig 3F), analyzed as previously described by us^51^. As expected, the IFN score markedly decreases over time from time-point A (earlier blood draw) to B (later one) in all patients and all cell types, corresponding to a decrease in viral loads between those time-points (Fig 3A, 3C). Notably, the decrease in IFN score between time-point A and B correlates strongly with the time difference between those two time-points (R=0.97, figure 3B), supporting the association between time and type-1 IFN signature. Symptom onset is reported to occur at a median of 5.2 days after infection^52^, and since blood draw A was taken at least 5 days after symptom onset (Fig 1D), at this time-point our patients would be expected to be on the descending slope of the viral load curve^53^. It explains the uniform decrease in viral load (and IFN score) between the two time-points. However, in two out of four progressive patients (and none of the six stable ones), both IFN score and viral load remained relatively high at time-point B. The gene expression signature of these two patients at time-point B (TP7B, TP8B) resembles the signature of other patients at the earlier time point A, while the other patients at time point B are closer to the healthy controls’ gene expression signature (Fig 3E). Although our sample size is not large enough to arrive at a definitive conclusion, this observation suggests that some progressive patients are slower in clearing the virus, possibly due to the immunosuppressive mechanisms noted above. Interestingly, the other two progressive patients (TP6B, TP9B) who had a low IFN score at the later time-point B and successfully cleared the virus, developed severe leukocytosis (>20,000/uL) with >80% neutrophils, which did not occur in any of the other COVID-19 patients.

### Tocilizumab effects differ across cell types and are associated with the levels of expression of *IL6R* and *IL6ST*

Eight of ten COVID-19 patients in our study were treated with tocilizumab, an anti-IL6 receptor (IL6R) antibody. As evident from Figure 3J, *IL6R* is highly expressed in monocytes, dendritic cells, neutrophils, CD4 T cells (including FoxP3 regulatory T cells (Tregs)) and naïve CD8 T cells. On the other hand, *IL6R* expression is low in the other types of lymphocytes including memory CD8, effector CD4 & CD8 T cells, gamma-delta T cells, B cells and NK cells. *IL6ST* (encoding gp130), responsible for signal transduction of IL-6 following binding to IL6R, is expressed in all types of PBMCs. To identify the transcriptional effects of tocilizumab treatment in COVID-19 patients, we compared gene expression changes from time point A to B for patients in the tocilizumab treatment group versus those not treated with tocilizumab (Fig 3D, Supp Fig S13). We highlight six tocilizumab responsive genes (*ARID5A, BCL3, PIM1, SOCS3, BATF, MYC*) that are associated with IL-6 pathway and known to be perturbed by tocilizumab treatment in rheumatoid arthritis patients^54^, indicating the consistent effect of tocilizumab in COVID-19 patients. Of note, those transcriptional changes by tocilizumab are observed mainly in the cell types that highly express *both IL6R* and *IL6ST*, such as naïve CD4 T cells, memory CD4 T cells, naïve CD8 T cells and Tregs. To quantify this effect, we generated an IL-6 score (a composite score for a pair of six tocilizumab responsive genes), and demonstrate a significant decrease of IL-6 score in CD4 T cells in all patients who received tocilizumab, except for one patient-TP7 (Fig 3K). Unlike the other progressive patients, this patient did not show an increase in plasma IL-6 levels, suggesting less contribution of IL-6 pathway in this patient’s pathophysiology.

Next, we sought to identify the other genes that are perturbed by tocilizumab in COVID-19 patients. To minimize the confounding effects of disease-related gene expression changes over time, we focused on genes that are not decreased over time in the non-tocilizumab group but significantly decreased following tocilizumab treatment (logFC>0.4, Supp Fig S17). We demonstrate that *S100A8* and *S100A9* expression are highly downregulated by tocilizumab treatment across the majority of the cell types, but not changed or even slightly increased in non-tocilizumab group, leading to a large logFC difference (Supp Fig S17). Given that a positive feedforward loop between S100A8/9 and IL-6 can drive pro-inflammatory circuit^55-57^ and that elevated serum S100A8/9 is one of hallmarks of severe COVID-19 patients^58,59^, it is possible that tocilizumab can exert its effect partly through the inhibition of S100A8/9 expression in COVID-19. Of interest, expression of *IL6R* is higher than that of *IL6ST* in myeloid cells, while it is lower in all other cell types, leading to a difference in *IL6R/IL6ST* ratio. According to a recent study^60^ this ratio determines the type of response to IL-6 signaling: anti-inflammatory classical signaling in cells with high *IL6R/IL6ST* (as observed in our myeloid cells) or pro-inflammatory trans-signaling in cells with low *IL6R/IL6ST* (non-myeloid cells), possibly explaining the observed difference between cell types in response to tocilizumab (Supp Fig S17). While we detected a response to tocilizumab at a cellular level, our study was neither designed nor powered to detect any clinical effect of the therapy.

### Surface protein-based immune-phenotyping of peripheral blood cells in COVID-19

We next constructed an independent immunophenotypic map of PBMCs using droplet-based CITE-seq^18^. To better identify cellular multiplets and enable us to super-load the cells onto the 10x platform, we used Cell Hashing technique and multiplexed 5-6 samples in each 10x reaction by using six hashing antibodies^61^. To integrate cellular surface protein and transcriptome measurements, we adopted 189 oligonucleotide-labeled antibodies (Total seq C antibody panel from BioLegend), which is the largest panel employed so far (Supp Table ST6). Seventeen out of 18 PBMC samples from COVID-19 patients were processed by CITE-seq with Cell Hashing. Antibody-derived tag (ADT) and Hashtag oligonucleotide (HTO) sequencing libraries were generated. CITE-seq samples were demultiplexed using hashing HTO counts (see Material and Methods section), which allowed for identification and removal of between-sample multiplets. Cells with unidentifiable sample origin were removed from the downstream analysis as well. Overall, 83.2% of the cells were retained (n=43,349; Supp Table ST7). Following unsupervised clustering, annotation for CITE-seq cells was performed with both gene expression and antibody-derived counts (ADT) by using a manually curated marker gene list (Supp Table ST8). We observed a cluster of undefined cells that are positive for multiple linage markers of ADT signals and/or display elevated signals unlikely to be explained by immunological evidence. Those cells (n= 8,032) were removed from the analysis, and all other cells were plotted on UMAP space (Fig 4A). As anticipated, both ADT and RNA measurements of marker proteins/genes demonstrate enrichment of CD3/*CD3E* within T cells; CD14/CD14 within the monocytic lineage; CD19/*CD19* within B cell lineage; CD56/*NCAM1* in NK cells; CD4/*CD4* in CD4^+^ T cells and monocytic linage to a lower degree; and CD8/*CD8A* enrichment within CD8^+^ T cells (Fig 4B). In order to investigate the coherence of both annotations, we calculated the percentage of shared cells between RNA and ADT annotated cell types (Fig 4C). Calculation of percentage shared cells between ADT and RNA based annotations showed reasonable overlap, confirming that cell type specifications are consistent robust in our analysis.

**Figure 4:**
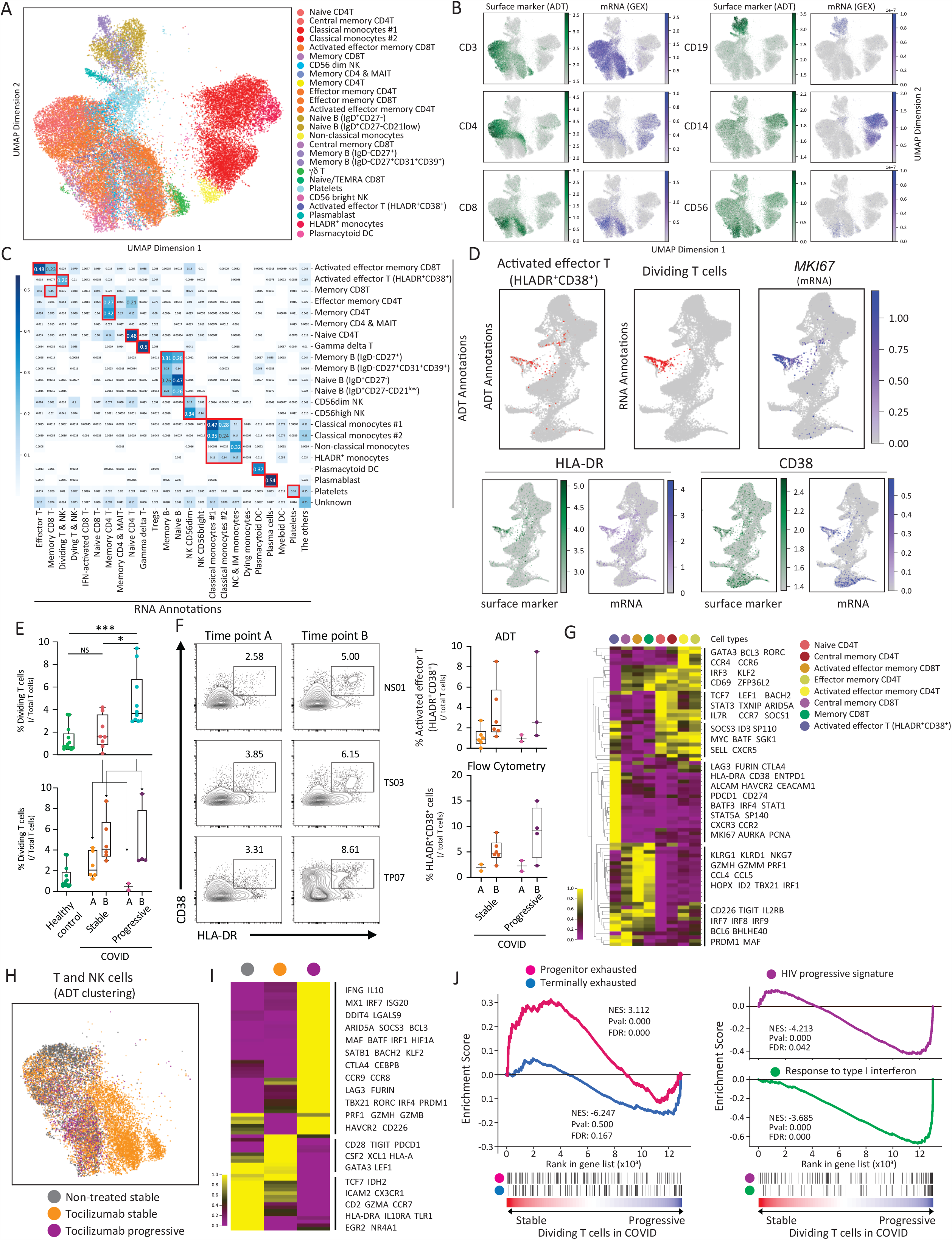
Multi-omics immune profiling identifies HLADR^+^CD38^+^ exhausted-like T cells in progressive COVID-19 patients **A** UMAP embedding of the ADT data from 43,349 cells across COVID-19 patients, excluding a cluster of non-specific staining (8,032 cells, see results section) **B** RNA expression overlay on single-cell ADT UMAP. ADT and mRNA expression for CD3, CD4, CD8, CD14, CD19, and CD56. **C** Heatmap showing the proportion of shared cells between ADT and RNA based annotations. Boxes highlighted in red show expected overlap between GEX and ADT. **D** Visualization of ADT cluster #15 (HLADR^+^CD38^+^ activated effector T cells) and TCR^+^ cells in GEX dividing T cells cluster on the GEX UMAP (red highlighted). ADT and RNA expression of *MKI67* (only RNA), HLA-DR/*HLA-DRA*, and CD38/*CD38* projected on GEX UMAP. **E** Box plots show frequency of dividing T cells within the total T cell population at early and late time-points (top), and in stable and progressive patients at early and late time-points (bottom). **F** Representative contour plot of flow cytometry analysis of PBMCs from COVID-19 patients (left). The frequency of HLADR^+^CD38^+^ cells assessed by ADT (top right) and flow cytometry (bottom right) in samples from stable and progressive COVID-19 patients is shown. **G** Heatmap showing the expression of curated genes across the different T cell clusters. **H** ADT UMAP of T and NK cells showing three severity and treatment groups by color. **I** Heatmap representation of DEGs across these three groups. **J** Left panel: GSEA of a signature of progenitor exhausted versus terminally exhausted CD8^+^ T cells in chronic infection (GSE84105). Right panel: A signature of HIV specific CD8+ T cells in progressive chronic HIV patients (GSE24081) and type-1 IFN response (GO:0034340). Progenitor exhausted signature (red), terminally exhaustion signature (blue), progressive HIV CD8+ T cell signature (purple), and type 1 IFN response signature (green) in the ranked list of genes differentially expressed by dividing T cells from stable versus progressive COVID-19 patients.

### HLADR^+^CD38^+^ activated T cells skew toward exhaustion-like phenotype in progressive COVID-19 patients

Among the overlapping cell types, we found that the ADT-annotated activated effector T cells cluster (HLADR^+^CD38^+^, ADT cluster #15) and the GEX-annotated dividing T/NK cluster (GEX cluster #20) represent a unique T cell subset (Fig 4C). Dual expression of HLADR and CD38 or higher expression of KI67 are known to mark a highly activated T cell population in acute viral infection^62-65^. We observed that *MKI67*-expressing T cells in the GEX dividing T/NK cluster were increased in COVID-19 patients, particularly in the late time point (Fig 4E). The population of ADT cluster #15 (higher HLADR and CD38) within T cells was also increased over time in COVID-19 patients. Further validation using flow cytometry with the same samples demonstrated identical results as determined by ADT-based calculations, confirming ADT successfully recapitulates the conventional flow cytometry-based protein signal (Fig 4F). These data strongly suggest that the temporal increase of this population is reproducible. The percentage of T cells overlapping between ADT cluster #15 and GEX dividing T/NK cluster was 26%, indicating expression of both HLADR^+^/CD38^+^ and KI67 marks a specific T cell subset in COVID-19 that increases during the disease course. This observation is supported by a recent study using flow cytometry with a larger number of COVID-19 patients^66^.

CITE-seq technology further allowed us to elucidate the transcriptional signature of this activated T cell population. Compared to the other T cell clusters (Fig 4G), the HLADR^+^CD38^+^ T cell cluster #15 exhibits enriched expression of co-inhibitory receptors (*LAG3, CTLA4, PDCD1, ENTPD1, HAVCR2*)^67,68^ and lower expression of naïve/stemness markers (*TCF7, LEF1*)^69-71^ and cytotoxic T cell markers (*NKG7, KLRG1, PRF1, GZMH*). Cell division markers (*MKI67, AURKA, PCNA*) are highly enriched in this cluster, consistent with the considerable overlap with the GEX dividing T/NK cluster. Transcription factors (TFs) promoting T cell exhaustion (*PRDM1, MAF*)^72^ are also enriched in this cluster. These data suggest that T cells in this cluster display skewed transcriptional signature toward T cell exhaustion.

Although our data demonstrated an increased frequency of HLADR^+^CD38^+^ T cells at the later time point in COVID-19 patients compared to controls, there was no clear difference in frequency between stable and progressive patients. We then determined whether there are transcriptional differences in this T cell cluster between stable and progressive COVID-19 patients. Progressive patients exhibited higher expression of type-1 IFN response genes (*MX1, IRF7, ISG20*) and cytotoxic/pro-inflammatory cytokines (*PRF1, GZMH, IFNG*), and lower expression of stemness/progenitor markers (*TCF7, LEF1*). Interestingly, while most of the co-inhibitory receptors were enriched in progressive patients (*LAG3, CTLA4, HAVCR2*), some were enriched in stable patients (*PDCD1, TIGIT*). Exhaustion driving TFs (*PRDM1, MAF*) and the immunoregulatory cytokine *IL10*, which is also co-expressed in exhausted T cells, were upregulated in progressive patients (Fig 4I). Recently, the heterogeneous features of exhausted T cells became appreciated and two major states are proposed: a progenitor or stem-like exhausted state^71,73^ and a terminally exhausted state^69,70^. Progenitor exhausted state is marked by higher expression of TCF7 and rich potential of producing cytotoxic cytokines, while terminally exhausted state is represented by high expression of multiple co-inhibitory receptors and dysfunctional features^69^. Our data demonstrated that the transcriptional characteristics of dividing T cells in progressive patients display a unique phenotype between progenitor exhausted and terminally exhausted states, which we call here a pre-terminally exhausted phenotype.

Gene set enrichment analysis (GSEA) further demonstrated that dividing T cells in the progressive COVID-19 patients exhibited more terminally exhausted T cell signature and type 1 IFN response signature than those in stable patients (Fig 4J, Supp Table ST9). Of note, this transcriptional signature in progressive COVID-19 patients overlapped with that of HIV specific T cells from HIV progressors compared to HIV controllers (Fig 4J). Given that the type-1 IFN pathway is enriched in terminally exhausted T cells in both tumor infiltrated T cells^69^ and chronic viral infections^74,75^, our data suggest that the stronger or prolonged type-1 IFN response in progressive COVID-19 patients may drive a phenotype of pre-terminal exhausted T cell signature.

LAG3 was the most upregulated co-inhibitory receptor in progressive COVID-19 patients, and its expression can be induced by IL-2 and IL-12 stimulation which are known to be associated with a Th1 signature^76^. LAG3 binds to MHC-II, LSECtin (encoded by *CLEC4G*)^77^ and FGL1^78^. Among the ligands for LAG3, we observed significant decreases of MHC-II molecules on myeloid cells and B cells in progressiveCOVID-19 patients, suggesting a possible decrease in LAG3-MHC-II interactions in progressive COVID-19 patients despite LAG3 being upregulated. Our differential connectomic analysis also highlighted the LAG3-MHC-II interaction in progressive versus stable COVID-19 patients (Fig 2C).

Taken together, our multi-omics analysis and flow cytometry-based validation revealed the increase of a unique T cell population marked by higher expression of HLADR, CD38, and KI67 in COVID-19 patients. Furthermore, transcriptional analysis of this population demonstrated a pre-terminally exhausted T cell signature in progressive COVID-19 patients with higher LAG3 expression. Reduced LAG3-MHC-II interactions between T cells and antigen-presenting cells may fail to complete the exhaustion program which is mainly driven by co-inhibitory receptor signals, resulting in aberrant expression of cytotoxic cytokines (Fig 4I) that may in turn contribute to immunopathology^30^.

### Skewed T cell receptor repertoire in CD8 T cells of progressive patient

In order to characterize the T cell receptor (TCR) repertoire relevant for immunity to SARS-CoV-2, we conducted a single-cell V(D)J analysis of COVID-19 samples. TCR data was captured for 44,876 cells in total with a median of 2,429 cells per sample. Quality assessment and control of the data filtered out 5,501 cells, leaving a median of 2,190 cells per sample. Based on these high-quality data, cells with the same V(D)J sequences of beta and alpha chains were grouped into clones. In total, 38,725 clones were identified with a median of 2,265 clones per sample (Fig 5A). The summary of the number of cells and clones and the cell type composition of these cells for each sample is shown in Figure 5A and 5B.

**Figure 5:**
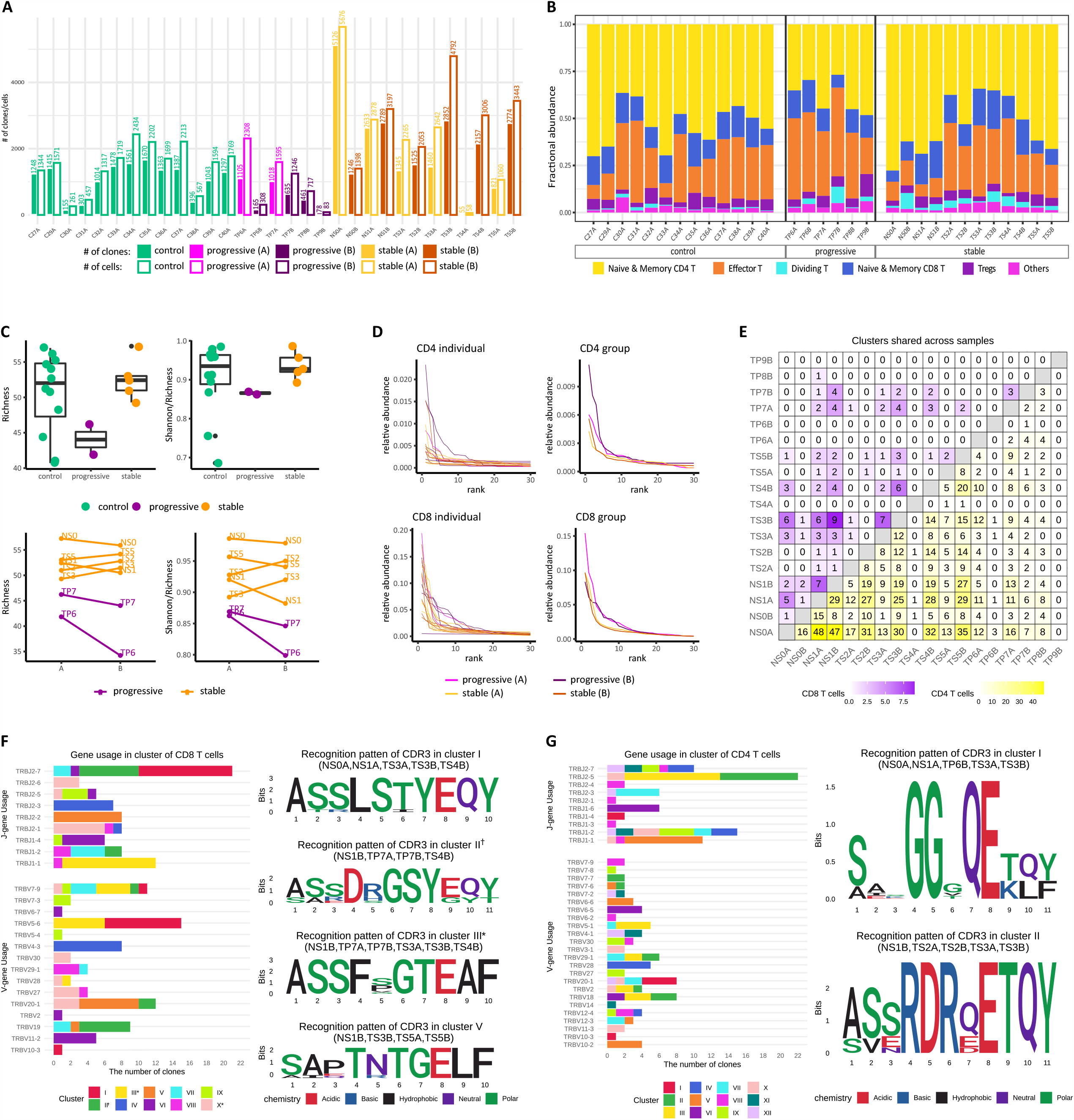
TCR data analysis on controls and COVID-19 patients. **A** The number of cells and clones across all samples. Cells with TCR data of low quality are excluded. **B** Fractional abundance of cells with high quality TCR data among different cell types. **C** Rarefied diversity indices (richness and evenness) of the naïve and memory CD8 T cells are different between stable and progressive patients presented in boxplots (top panels). Direction of changes in these diversity indices after treatment is different between progressive and treated stable patients (bottom panels). **D** Rarefied relative abundance of the top 30 clones from CD4 T cells (top panels) and CD8 T cells (bottom panels) shows different expansion rate distribution between stable and progressive patients. The expansion rate of CD8 T cells is also much higher than that of CD4 T cells. **E** The number of clone clusters identified by GLIPH2 in CD4 T cells (lower triangle) and CD8 T cells (upper triangle) that have clones from every pair of samples based on the top 24 CD8 and 172 CD4 SARS-CoV-2 specific clone clusters. **F** Clone clusters’ TRBV and TRBJ gene usage distribution in CD8 T cells based on the top 10 SARS-CoV-2 specific expanded clone clusters. The CDR3β motif found in each of the chosen 4 clusters with global similarity is shown as well as the samples that contribute clones to the cluster. * Clusters with dividing CD8 T cells. † Clusters with IFN-activated CD8 T cells. **G** Clone clusters’ TRBV and TRBJ gene usage distribution in CD4 T cells based on the top 12 SARS-CoV-2 specific expanded clone clusters. The CDR3β motif found in the top 2 clusters is shown as well as samples that contribute clones to them.

Alpha diversity with rarefaction was calculated using the Alakazam R package^79^ for both memory and naïve CD4 T cells and memory and naïve CD8 T cells. The diversity of memory and naïve CD8 T cells at both time points shows much lower richness and evenness (i.e. Shannon index/richness) in progressive patients than stable patients (Fig 5C), which is consistent with higher expansion of CD8 T cell clones in progressive patients (Fig 5D). This difference in alpha diversity was not observed in memory and naïve CD4 T cells. In addition, progressive patients have a further decrease in CD8 clonal richness and evenness at the later time-point B (Fig 5C, bottom panel).

### Identification of COVID19-specific CDR3 regions

In order to identify characteristics of the TCR regions that may confer specificity to SARS-CoV-2, we used GLIPH2^80^ to assess similarity of complementarity-determining region 3 (CDR3) sequences among COVID-19 patients. We specifically looked for CDR3 motifs in β chains that were shared across several COVID-19 patients but in none of the 13 control subjects. Stringent filters were applied to the GLIPH2 CDR3 specificity groups (or clusters) to improve accuracy, including requiring Fisher’s score < 0.0001, ≥3 unique TCRs in the specificity group and significant V-gene bias (P < 0.05). The filtered specificity groups with any clone from control samples were filtered out to enhance the likelihood of specificity to SARS-CoV-2 instead of to other common viruses such as cytomegalovirus. After heavy filtering, 24 and 172 groups remained for CD8 and CD4 T cells, respectively. Most of the identified specificity groups included clones from different samples, suggesting a large similarity in the CDR3 sequence in the potential SARS-CoV-2 specific clones (Fig 5E). To further enhance the specificity to clonally-expanded SARS-CoV-2 responsive T cells, we focused on 10 CD8 and 12 CD4 groups that have clones from ≥3 subjects with at least one of these clones with ≥2 cells. The V and J gene usage analysis suggested a dominant usage of some VJ combinations such as TRAV5/TRAJ12/TRBJ2-7/TRBV5-6 for cluster 1 in CD8 T cells (Fig 5F, Supp Fig S15). For the CD4 T groups, there is no obvious V gene usage bias and the J gene usage is dominated by TRBJ2-5 (Fig 5F-G).

Among the 10 and 12 SARS-CoV-2 specific and expanded groups, we further chose those that include clones from ≥3 different COVID-19 patients with ≥55% clones having more than one cell, resulting in five and two groups for CD8 and CD4 T cells, respectively. The chosen clusters are also the top five and two clone clusters with the best composition score by GLIPH2, which measures the strength of a specificity group based on global/local similarities, enrichment of common V-genes, enrichment of a limited CDR3 length distribution, enrichment of clonally expanded clones, and cluster size. This suggests that the chosen specificity groups are likely from SARS-CoV-2 specific expanded clones, are shared across COVID-19 patients and have a highly conserved CDR3 amino acid sequence. All specificity groups are identified based on global similarities in CDR3 region, except for cluster IV in CD8 T cells, whose member clones have different CDR3 lengths but share the motif “QDIG”. The CDR3 sequence motifs of the specificity groups with global similarity are shown in Figure 5F and 5G.

### Single-cell V(D)J B cell receptor repertoire analysis

For each sample in the single-cell V(D)J library, a summary of the number of cells, frequency of each B cell type (naive B, memory B, and plasma cells), and frequency of each isotype (IGHM, IGHD, IGHG, and IGHA) is provided in Figure 6 A, B, and C, respectively. Overall, the single-cell V(D)J library contains 7,177 cells distributed across 18 samples.

**Figure 6:**
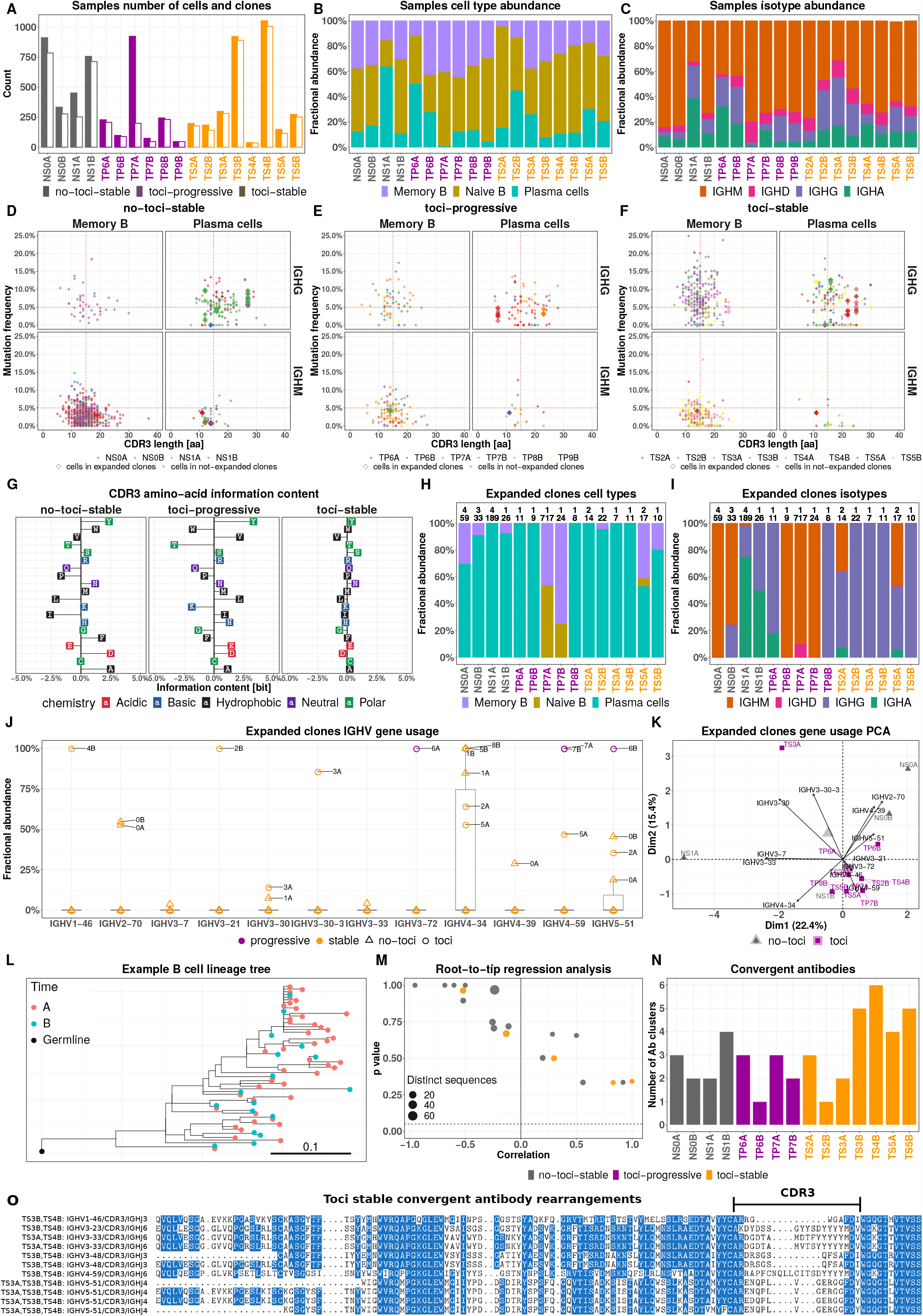
BCR data analysis. **A** Number of cells (closed-bars) and number of clones (open-bars) in stable patients with no treatment (grey), progressive patients under treatment (purple), and stable patients under treatment (orange). **B** Fractional abundance of memory B cells (purple), naive B cells (brown), and plasma cells (purples) in each sample. **C** Fractional abundance of IGHA (green), IGHD (pink), IGHG (purple), and IGHM (green) cells in each sample. **D, E, and F** CDR3 amino acid length (x-axis) and mutation frequency (y-axis) for each cell type (memory B cell and plasma cell columns) and isotype (IGHG and IGHM rows) of stable patients with no treatment (D), progressive patients under treatment (E), and stable patients under treatment (F). Point colors indicate different samples in each compartment. Vertical dashed line represents 15 amino acid CDR3 length reference point and horizontal line represents 5% mutation frequency reference point. Points with larger size belong to the expanded clones. **G** CDR3 amino acid usage difference comparing time-points B versus A in stable patients with no treatment, progressive patients under treatment, and stable patients under treatment. **H** Fractional abundance of expanded clones memory B cells (purple), naive B cells (brown), and plasma cells (purple) in each sample. Labels in each bar represent the number of expanded clones (top) and number of cells (bottom). **I** Fractional abundance of expanded clones IGHA (green), IGHD (pink), IGHG (purple), and IGHM (green) in each sample. Labels in each bar represent the number of expanded clones (top) and number of cells (bottom). **J** IGHV gene (x-axis) fractional abundance (y-axis) of expanded clones in each sample (label). The color of each point represents the status of the disease (purple for progressive and orange for stable patients) and the shape of each point represents the treatment (triangle for patients without treatment and circle for patients under treatment). **K** Principle component analysis (PCA) of IGHV genes (arrows label) based on the fractional abundance from expanded clones of each sample (points label). Colors and shapes in panel K represent patients grouped based on treatment (grey-triangle for patients without treatment and purple-square for patients under treatment). **L** An example of a B cell clonal lineage tree. Branch lengths represent the expected number of substitutions per codon (see scale bar). **M** A root-to-tip correlation analysis. Pearson correlation coefficient between divergence and time within each B cell lineage tree (x-axis), with corresponding p values calculated using a permutation test (y-axis). The size of each point corresponds to the number of distinct sequence/time point combinations within each clone. Dashed line shows p-value = 0.05. **N** shows the number of convergent antibody clusters within each sample. **O** Convergent antibodies (VDJ) that are specific to patients with stable status and under treatment. The labels represent V and J genes for each given antibody. The complementarity determining region-3 (CDR3) regions are shown with arrowed-line.

### COVID-19 patients with stable status show higher mutation frequency and longer CDR-H3 length

IGHV/IGHJ mutation frequency and CDR-H3 length varied by antibody isotype and cell type within COVID-19 patients (Fig 6 D-F). IGHM memory and plasma B cells had IGHV/IGHJ mutation frequencies significantly lower than 5% (mutations/nucleotide, p-value < 0.01), regardless of treatment or disease progression group. As expected, IGHG memory B cells and plasma cells had IGHV/IGHJ mutation frequencies higher than IGHM cells. In particular, plasma cells in stable COVID-19 patients without treatment had mutation frequencies significantly higher than 5% (mean = 5.6+/-3%; p-value < 0.01). Memory cells in stable patients under tocilizumab treatment had even higher mutation frequencies (mean = 7.5+/-4.6%).

The CDR-H3 region length of IGHG and IGHM B cells generally varied between 10 to 20 amino acids (we used 15 amino acids as a reference point for downstream CDR-H3 length comparisons) across all cell types (Fig6D-F). However, the CDR-H3 length of IGHG plasma cells in stable COVID-19 patients was significantly larger than 15 amino acids (p-value < 0.01), while the CDR-H3 length of IGHG memory cells do not show significant differences from 15 amino acids (mean = 15.2+/-3.9 amino acids across all samples). On average, CDR-H3 lengths of IGHG plasma cells were larger in stable no-tociluzimab patients (mean = 18.4+/-5.4 amino acids) than in stable tociluzimab-treated patients (mean = 17.2+/-5 amino acids).

### Stable patients under treatment do not show change in CDR-H3 amino acid usage

We sought to investigate the differences in CDR-H3 amino acid usage between the two blood draw time points (A and B) (Fig 6G). We tackled this query by calculating the conditional information content of each amino acid in the CDR-H3 segment at time point B with respect to time point A. We averaged the conditional information contents for patients belonging to three different groups: (1) stable patients under no treatment (no-tocilizumab-stable); (2) progressive patients under tocilizumab treatment (tocilizumab-progressive); and (3) stable patients under tocilizumab treatment (tocilizumab-stable). Our results indicate that the profile of amino acid usage for tocilizumab-stable patients is quite different from the other groups. In fact, the IGH repertoires of tocilizumab-stable patients do not show any change in preferences toward usage of specific amino acids in their CDR-H3 segment between the time-points. In contrast, the information content profiles of tocilizumab-progressive and no-tocilizumab-stable patients vary across amino acids. In particular, tocilizumab-progressive and no-tocilizumab-stable patients show evidence of increased usage of alanine (A), aspartic acid (D), and tyrosine (Y), and decreased usage of proline (P), glutamine (Q), and threonine (T) at time-point B relative to A.

### High frequency of plasma cells in all patients, IGHG in tocilizumab-stable patients, and IGHM in tocilizumab-progressive patients within expanded clonal lineages

To explore antigen-driven B cell responses in the COVID-19 patients, we investigate expanded clonal lineages (Fig 6H-I). We identified 20 expanded clones with members found in 15/18 samples containing 1157/7177 cells (16% of all B cells). Within this group, 5 clones belonged to patient NS0, 4 belonged to patient NS1, 3 belonged to patient TS2, 1 belonged to patient TS3, 1 belonged to patient TS4, 2 belonged to patient TS5, 2 belonged to patient TP6, 1 belonged to patient TP7, and 1 belonged to patient TP8. Features that were significantly enriched in expanded clones were determined by comparing their fractional abundance in expanded clones to those in non-expanded clones. Plasma cells were significantly enriched in the expanded clones (mean = 78% across all samples; mean odds ratio = 5.7; p-value < 0.01) (Fig 6H). Expanded clones in samples TP7A and TP7B did not contain any plasma cells. IGHG cells were significantly enriched in expanded clones from tocilizumab-stable patients (mean = 84% across 6 samples, mean odds ratio = 8.2; p-value < 0.01). IGHM cells were enriched in expanded clones from tocilizumab-progressive patients (mean = 58% across 5 samples, mean odds ratio = 1.2) (Fig 6I).

We further sought to investigate the mutation frequency and CDR-H3 length of cells within expanded clones. We observed that the mutation frequency of IGHG plasma cells from stable COVID-19 patients was higher on average (mean = 5.5+/-4%) than that in progressive patients (mean = 3.2+/-0%), regardless of treatment status. We further observed that the CDR-H3 length of IGHG plasma cells from stable COVID-19 patients was significantly larger than 15 amino acids (p-value < 0.01). In particular, patients without treatment had a larger mean CDR-H3 length (mean = 20.2+/-6.6 amino acids) than those under treatment (mean = 17.5+/-5.4 amino acids). IGHG plasma cells from progressive COVID-19 patients had CDR-H3 lengths significantly shorter than 15 amino acids (mean = 14.5+/-8.2 amino acids; p-value < 0.01).

Selection of particular IGHV genes in response to a particular antigen has been observed in other antiviral responses, such as the preference for IGHV1-69 in response to some influenza virus antigens^81,82^. Therefore, we sought to identify IGHV genes under selection in COVID-19 patients (Fig 6J). We identified that IGHV4-34 gene is highly used in expanded clones of stable patients with odds ratio of ∼10 among patients under treatment and ∼9.5 among patients without treatment. tocilizumab-progressive patients showed a lower usage of IGHV4-34 with an odds ratio of ∼5.9. We further performed principal component analysis (PCA) of IGHV gene usages in expanded B cell clones (Fig 6K). We have identified a cluster of patients under treatment, including both stable and progressive, whose corresponding expanded clones only contain IGHV1-46 (100% in TS4B), IGHV3-21 (100% in TS2B), IGHV3-30-3 (85% in TS3A), and IGHV3-72 (100% in TP6A).

### COVID-19 patients show a high level of unmutated IGHG clones, and large clones with stable SHM frequency between time points

BCR sequence analysis can provide important information about the dynamics of the B cell response within COVID-19. For example, prior work (Nielsen et al. 2020) has demonstrated an elevated proportion of unmutated (median SHM < 1%) IGHG B cell clones in COVID-19 patients compared to healthy controls. This could be an indication of early class switching before GC entry in a primary immune response. We similarly observed that between 0% and 45.7% of IGHG clones within each patient were unmutated (mean = 19.8%; Fig 6D, S16), considering both expanded and non-expanded clones. Further, tocilizumab-stable patients had a higher fraction of unmutated IGHG clones compared to tocilizumab-progressive patients (mean = 30.8% vs 11.3%, respectively), though this difference was not significant (Fig S16, p-value = 0.057, Wilcoxon test). These clones were primarily composed of memory and plasma cells, in contrast to IGHM clones, which were primarily composed of IGHM cells (Figure S16). We also observed multiple diverse B cell clones, both expanded and non-expanded, spanning both timepoints. To characterize potential affinity maturation in these clones, we built B cell phylogenetic trees for all clones containing at least three sequences that were either distinct or found at different timepoints (largest shown in Figure 6L). We observed a relatively high level of SHM in these clones at timepoint A (mean = 4.7%). We then used a phylogenetic root-to-tip correlation test (Methods) to determine if divergence from the germline sequence increased between timepoints A and B in these 20 clones. None showed a significant positive correlation between sample time and divergence from the sequences’ most recent common ancestor (i.e. p-value > 0.05; Fig 6M). These results indicate a lack of measurable SHM accumulation between time-points in these clones.

### Convergent antibody rearrangements are elicited in COVID-19 patients

We identified 19 convergent antibody clusters across eight out of ten patients. 79% (15/19) of convergent clusters included antibodies from tocilizumab-stable patients, 47% (9/19) included antibodies from no-tocilizumab-Stable patients, and 31% (6/19) of convergent clusters included antibodies from tocilizumab-progressive patients (Fig 6N). We next focused on the tocilizumab-stable patients, which were represented in the most convergent clusters. We identified six convergent clusters which were composed only of tocilizumab-stable patients (Fig 6O). These convergent antibody clusters spanned between two patients, and were composed of IGHV1-46/IGHJ3, IGHV3-23/IGHJ6, IGHV3-33/IGHJ6, IGHV3-48/IGHJ3, IGHV4-59/IGHJ6, IGHV5-51/IGHJ4 gene rearrangements.

## Discussion

Although initial studies in COVID-19 patients with severe respiratory failure revealed a dysregulated immune system with hyper-inflammatory responses and lymphopenia^15-17,83-85^, there are critical deficits in our understanding of the mechanisms underlying this immune dysfunction. Here, we applied multimodal single-cell analysis using 5’ scRNA-seq, CITE-seq, TCR, and BCR sequencing on 18 samples from 10 patients with COVID-19, which were compared to 13 control samples. Our experimental design allowed us to determine: 1) the dynamics of the immune response in COVID-19 over time; 2) general immune cell features in COVID-19 patients; 3) the specific immune signature associated with progressive disease; 4) an in-depth exploration of adaptive immunity using T cell and B cell repertoire analysis in COVID-19; and 5) the effects of tociluzimab treatment. Our unbiased systems biology approach utilizing novel multimodal single-cell analysis techniques reveals the temporal dynamics of immune responses to this disease. It highlights the unique immune features that distinguish stable and progressive COVID-19 patients. The immunological networks characterized in our study improve the understanding of the abundant cellular interactions and effects that promote pathology in severe disease, including dyssynchrony of the innate and adaptive immune response with anti-inflammatory and with unique “pre-exhaustion” immune features.

We observed dominant effects of a type-1 IFN response across all immune cells in all COVID-19 patients, especially at the earlier time-point A, consistent with the acute viral infection^86^. In this regard, we highlight the IFN-activated CD8^+^ T cell cluster as the extreme archetype of type-1 IFN response in T cells, which was almost exclusively found in COVID-19 patients, especially at time-point A (Figure 1E, 1I). Type-1 IFN response was the main driver of gene expression changes between progressive and stable subjects, as depicted in Figure 2D. Progressive patients had higher expression of ISGs in all major cell types. Type-1 IFN response, as reflected by the IFN gene score, decreased overwhelmingly from time point A to B in all patients, and highly correlated with time (R=0.97, Fig 3A-B). This is not surprising given that the first blood draw was obtained at least 5 days after the beginning of symptoms, the onset of which occur a median of 5.2 days after infection^52^. At this time-point, patients are expected to be on the descending slope of the viral load curve^53^, which type-1 IFN response closely follows (R=0.8 for the correlation between log10 viral load and IFN score). Interestingly, the ISG signature at the later time-point B returned towards control level in all patients except for two out of four in the progressive group (Fig 3E), in which both IFN score and viral load remained relatively high. This is consistent with a previous study in which severe COVID-19 patients had a higher viral load and longer virus shedding than mild cases^87^.

The joint profiling of gene expression and surface proteins shed light on a specific T cell population that expressed higher HLADR, CD38, and cell cycle markers (*MKI67, PCNA, AURKA*) and expanded with time. Although there was no significant difference in frequency noted between progressive vs stable COVID-19 patients in our cohort, we demonstrated the enrichment of ISGs and unique set of co-inhibitory receptors (LAG3 and TIM3, coded by *HAVCR2*) in progressive patients. Interestingly, TIGIT and PD1 (coded by *PDCD1*) were relatively higher in stable patients together with *TCF7* and *LEF1*, the markers for progenitor exhausted T cells^69^. We label this intermediate state of exhaustion as “pre-terminal” exhaustion. In general, T cell exhaustion is observed in chronic infectious diseases and cancer where T cells receive sustained stimuli in an immunoregulatory milieu^88,89^. In contrast, our observation in severe SARS-CoV-2 infection indicates that a pre-terminally exhaustion phenotype can also be induced in acute viral infection^90^, which is not commonly seen. Chronic TCR stimulation is necessary to induce terminally exhausted T cells. However, COVID-19 patients in our study presumably have not been exposed to antigens for a long enough time period to drive an exhaustion program, suggesting there may be other factors facilitating the T cell exhaustion program in severe COVID-19 cases. Among signatures observed in progressive patients, elevated *LAG3* expression was detected across most T/NK cell subsets, except for the Treg cluster. Given that the expression of MHC-II molecules (which are the ligands for LAG3) is markedly downregulated in APCs in progressive COVID-19 patients, the disruption of LAG3-MHC-II interaction might play a critical role in COVID-19 immunopathology. While the trigger inducing LAG3 on T cells is not well understood, co-expression of ISGs in T cells from progressive patients suggests a possible role of TYPE-1 IFN on LAG3 expression. Further studies focusing on the molecular mechanisms by which SARS-CoV-2 induces LAG3 on T cells are warranted.

A skewed phenotype of myeloid cells toward a regulatory/immunosuppressive signature has been previously reported in severe COVID-19^16,17^. In contrast, a pro-inflammatory monocyte phenotype has been shown by others in patients with severe COVID-19^83-85^. Our findings of an anti-inflammatory monocyte cluster that is increased in COVID-19 patients, along with suppressive/tissue-repair gene expression changes in monocytes that are accentuated in progressive patients (including *CD163, IL1R2, AREG, MRC1, HAVCR2, LGALS9, IL10*), support the former evidence. We also observed the down-regulation of IL-1β mRNA expression in COVID-19 patients, indicating a shift from pro-inflammatory to a regulatory phenotype in monocytes in COVID-19. Our multimodal single-cell analysis demonstrated that these regulatory monocytes are transcriptionally distinguished from the other classical monocytes and resemble myeloid-derived suppressor cells (MDSCs), a heterogeneous myeloid cell population characterized by strong immunosuppressive function increased in chronic infection and cancer. The increased fraction of MDSC-like anti-inflammatory monocytes facilitates the resolution of inflammation during acute viral infection, transitioning from inflammation to tissue repair^91,92^. However, an inadequate anti-inflammatory/tissue repair response can delay virus clearance, cause chronic inflammation, and lead to excessive tissue damage and even tissue fibrosis^36,93-95^. Therefore, a regulated transition of the immune response from inflammation to tissue repair with appropriate kinetics is essential to restore tissue homeostasis.

The platelet gene signature in monocytes, which indicates the existence of platelet-monocyte complexes (or aggregates), was markedly increased in progressive patients (Fig 2L). A recent study suggested that serum IL-6 and IL-8 levels were significantly elevated in COVID-19 patients^96^, and these cytokines can activate platelets efficiently^97^. Activated platelets have a capacity to induce anti-inflammatory macrophage polarization, and this increase in platelet signature might amplify IL-10 production^39,98^. While IL-10 is critical to protect the host from tissue damage during acute immune responses, it also exhibits a detrimental immunopathogenic effect during acute viral infections by downregulation of MHC-II expression^36^. Our data clearly demonstrated the upregulation of IL-10 and the downregulation of MHC-II expression in monocytes which may contribute to the detrimental clinical course in progressive patients compared with stable ones.

Our T cell receptor repertoire analysis shows a higher expansion/dominance and a lower richness of CD8 but not CD4 T cell clones in progressive COVID-19 in progressive patients relative to stable patients. In an attempt to identify specific clonotypes that are relevant to T cell response against SARS-CoV-2, we studied the CDR3 motifs that are shared by several COVID-19 patients but absent from the control subjects. Using this approach, we identified 10 CD8 and 12 CD4 specificity groups, and described their specific V & J gene usage patterns. Moreover, we describe the specific CDR3 motifs that have the highest likelihood of being COVID-19 specific. In future experiments, these TCRs will be investigated for antigen specificity.

During viral infection, B cells are critical for the production of protective antibodies. Establishment of a diverse repertoire of antibodies is imperative to protect a host from pathogens, as well as to generate effective immune responses. One key finding is that CDR-H3 amino acid usage profiles were highly variable among patients (Fig 6G). In particular, there was no preference for any amino acid between early and late time points in stable patients who received tocilizumab. In contrast, progressive and stable patients who were not treated with tocilizumab showed a different profile of CDR-H3 amino acid selection which generally varied across amino acids with a trend toward increased usage of alanine (A), aspartic acid (D), and tyrosine (Y) at time-point B relative to A. The importance of CDR-H3-tyrosine for optimal antibody binding was previously shown for influenza A virus^99^. Another finding focused on the detection of convergent antibodies with highly similar VDJs across COVID-19 patients (Fig 6N, 6O).

Our V(D)J B cell receptor repertoire analysis further suggests a complex B cell response in COVID-19. Consistent with the expectations of a primary immune response, we observed an high proportion of unmutated IGHG B cell clones, which has been reported in at least one other analysis of BCR repertoires from COVID-19 patients^100^. However, we also observed multiple mutated B cell clones that were persistent across the two measures timepoints, but did not measurably accumulate SHM between timepoints (Fig 6L, 6M)^101^. These could result from cross-reactivity of memory B cells with other common corona-viruses, which has been documented in T cells^86^. Such memory B cells would have already accumulated SHM and likely avoid germinal-center re-entry^102^. This scenario may account for the quick expansion of plasma cells in COVID-19 patients (Fig 1E), which has also been reported by others^17,66^. It is also possible these are clones are non-coronavirus specific persistent clones sometimes observed in healthy older patients^103^. Importantly, these analyses were performed over a short time interval, and used a relatively small number of B cells with unknown specificity. We note, however, that the lack of observable SHM increase between timepoints is consistent with another recent study which found that levels of SHM in SARS-CoV-2 specific antibodies were stable (∼3%) between 8 and 42 days post-diagnosis^104^.

The relatively small sample size (18 COVID-19 samples and 13 controls) is a limitation of this study, especially for the analysis of certain subgroups (e.g. only four samples from patients who did not receive tocilizumab). However, it is larger than most COVID-19 scRNAseq studies published to-date^17,83,84,96^, and the similarity of baseline characteristics between stable and progressive patients and in comparison, to controls (Table 1) helps increase confidence in our results. Although the timing of blood draw A (time-point A) relative to hospitalization was consistent across subjects, the timing of blood draw B (time-point B) was variable. We mitigated that by taking into account the variable time span between the two blood draws in some of the analyses, e.g. for the analysis of IFN score changes over time shown in Fig 3A & B. This unique exploration of gene expression changes over time adds an essential dynamic layer that is critical to understand the biology of an acute viral disease. Lastly, our analysis mostly relied on RNA-based analyses including gene expression and TCR/BCR repertoire analysis, with some protein-level validation by CITE-seq and flow cytometry. Additional mechanistic validation, while beyond the scope of this study, is warranted in future studies.

In conclusion, our in-depth multi-omics assessment of peripheral immune cells at single-cell resolution across patient severities and time highlights the desynchronized adaptive and innate immune response in progressive COVID-19 patients. A prominent type-1 interferon response is observed across all immune cells, especially in progressive patients, and wanes over time in correlation to the decrease in viral loads. Anti-inflammatory monocyte response and a pre-exhaustion phenotype in activated HLADR^+^CD38^+^ T cells are hallmarks of progressive disease. Skewed T cell receptor repertories in CD8 T cell and uniquely enriched V(D)J sequences are identified in COVID-19 patients. B cell receptor repertoire analysis reveals a high level of IGHG B cell clones with little or no somatic hypermutation, consistent with a primary immune response, as well as mutated clones which may reflect stimulation of pre-existing memory B cells. Overall, our comprehensive immune profiling underscores the desynchronized innate and adaptive immune interaction in progressive COVID-19, which may lead to delayed virus clearance. This high-resolution understanding of the immune cell profiles underlying severe COVID-19 will enhance our ability to develop immunomodulatory therapeutic approaches to prevent progression in COVID-19 patients.

## Data Availability

Our data will be publicly available on GEO and the results can be explored through the COVID-19 Cell Atlas Data Mining Site (www.covidcellatlas.com).

## Acknowledgements

We would like to thank Yale Environmental Health and Safety (EHS) office, particularly Dr. Maren Schniederberend, for providing the safety guidance for working with COVID-19 samples and appreciate the helpful discussions we had with Drs. Michela Comi and Khadir Raddassi. Above all we wish to thank our families; your support was paramount to our success in this herculean project.

## Author Contributions

<u>Overall study design;</u> A.U., T.S.S., C.D.C., D.A.H., and N.K.

<u>Biospecimen collection/processing;</u> A.U., T.S.S., J.C.S., H.A., G.W., G.D., C.C., P.W., M.M., the Yale IMPACT research team.

<u>Epidemiological and clinical data collection;</u> A.U., P.W., J.F., S.B., L.S., C.B.F.V., A.L.W., N.D.G., M.M., the Yale IMPACT research team, A.I.K., S.F.F., A.I., A.C.S., C.D.C.

<u>Data analysis;</u> A.U., T.S.S., N.N., X.Y., A.Y.Z., V.G., J.C.S., H.A., Y.L., C.C.Jr., W.D., M.C., M.S.B.R., K.H., G.W., Z.W., N.G.R., N.L., A.C., A.M., H.M., D.V.D., H.Z., S.H.K., L.E.N., R.R.M.

<u>Original draft writing;</u> A.U., T.S.S., N.N., X.Y., V.G., H.A., W.D., M.C., K.H., H.Z., S.H.K., D.A.H., N.K., C.D.C.

<u>Reviewing and editing the manuscript;</u> All authors participated in editing the manuscript.

<u>Supervising the study;</u> D.A.H., N.K., C.D.C.

## Supplementary materials

### Methods

### Supplementary figures

S1 Heatmap of main markers used for manual cell type annotation

S2 UMAP of PBMC with Seurat clusters

S3 UMAP with SingleR automated annotation

S4 Comparison of the two sample processing methodologies: CITE-seq and conventional (non-CITE)

S5 Top 20 DEGs in COVID-19 vs controls, according to cell type

S6 AREG expression

S7 Expression of MHC-II molecules in stable and progressive COVID-19 patients vs controls

S8 Relative cell counts of the monocyte clusters and HLA-DR expression

S9 *IL10* expression in progressive vs stable patients, and correlation with IFN score

S10 UMAP of monocytes (sub-clustered)

S11 *IL1B* and *TNF* expression in monocytes

S12 UMAP of CD8 T cells (sub-clustered)

S13 Analysis of the effects of tocilizumab

S14 CITE-seq analysis-supplementary figure

S15 TCRα gene usage in CD8 T cells that belong to COVID19-specific CDR3 clusters

S16 Unmutated B cell clone analysis

### Supplementary tables

ST1 Patient baseline characteristics

ST2 DEGs in monocytes of progressive vs stable COVID-19 patients

ST3 DEGs in CD4 T cells of progressive vs stable COVID-19 patients

ST4 DEGs in CD8 T cells of progressive vs stable COVID-19 patients

ST5 DEGs in B cells of progressive vs stable COVID-19 patients

ST6 TotalSeq-C human Panel list that was used for CITE-seq

ST7 CITE-seq de-hashing statistics and barcodes

ST8 CITE-seq Panel for annotation ST9 GSEA custom gene sets

ST10 Ligand-receptor pairs for the connectome analysis

